# The Survival Analysis of Veterans’ Administration Lung Cancer Dataset

**DOI:** 10.1101/2022.05.22.22275430

**Authors:** Ewuru Deborah Amaka, İlker Etikan

**Affiliations:** Near East University, North Cyprus. Faculty of Medicine, Department of Biostatistics.; Near East University, North Cyprus. HOD, Department of Biostatistics. Faculty of Medicine

**Keywords:** Survival Analysis, Lung cancer, Kaplan-Meier, Life table, Cox Regression, Exp (β)

## Abstract

**AIM:** This study will describe the notion about survival analysis and the variability between diverse methods and test statistics used in analyzing survival data.

**METHODS:** This study will be using the Veterans’ Administrative Lung Cancer dataset(R Dataset) of 137 patients having advanced inoperable Lung cancer, carried out by the US Veterans Administration. Out of 137 male patients, 69 patients received the Standard test while 68 patients received Chemotherapy test and 64 patients died in each treatment group while only 9 patients were alive (censored) at the end of the study. This dataset will be analyzed using the SPSS version to determine whether there is a statistically remarkable difference between the survival distributions of the cell types of Lung cancer using Life tables, and also between the Standard test and Chemotherapy test treatment groups using Kaplan Meier method. Various covariates were also analyzed to find out if they have any effect on the death of the male patients with advanced inoperable lung cancer using Cox-Regression.

**RESULTS:** The Wilcoxon (Gehan) statistic from Life table method shows a p-value of 0.000 less than the 95% confidence level indicating that there is a statistical significance between the Lung cancer cell types. The three statistical tests of Kaplan-Meier method which include Log Rank (Mantel-Cox) test, Breslow (Generalized Wilcoxon) and Tarone Ware tests showed that the survival distribution between the two Lung cancer treatment groups are equal using an alpha level of 0.05.The Standard test and the Chemotherapy test drug treatment show the same effect on the survival time of the patients, no treatment is more beneficial than the other. Cox Regression result showed that Squamous cell type2(4), Adeno-carcinoma cell type (1), Small cell type (3) and karnofsky score are statistically significant while Large cell type2 (2), prior therapy, age, diagnosis time and the treatment group were found not significant.

**CONCLUSION:** The treatment difference does not show any remarkable effect on the survival time of male patients with Lung Cancer.

## INTRODUCTION

Survival Analysis is concerned with time to event data. It measures the time a person or group of people enters a study and when they experience an occurrence of interest. In survival analysis, the precise beginning and ending dates are not important because a subject can join a study at any moment. It is important to note that the subjects under observation may not experience any event until the end of the study because of difficulty in observing all the participants that dropped out, unavailability of post-treatment data for the verification of the occurrence or non-occurrence throughout the follow-up time is known as **CENSORED DATA**. Some statistical analysis techniques remove censored data from the analysis while in Survival analysis they are handled without processing them as incomplete or missing data. It also allows the researchers to observe and compare these data. Observed data of subjects that enter a study at different times and varied levels of exposure are gradually censored. Subjects that their event of interest did not occur at the end of the follow up are **Right Censored** whereas the subjects that their birth has not been seen are **Left censored**.

This study will be using the Veterans’ Administrative Lung Cancer dataset (R Dataset) of 137 patients having advanced inoperable Lung cancer, carried out by the US Veterans Administration. Out of 137 male patients, 69 patients received the Standard test while 68 patients received Chemotherapy test and 64 patients died in each treatment group while only 9 patients were alive (censored) at the end of the study. This dataset will be analyzed using the SPSS version. This dataset contains nine variables which include prior therapy (whether a patient received treatment before the recent one, 0 indicates no while 10 indicates yes),age of patients in years, diagnosis time(time between diagnosis and the commence of the study), karnofsky performance score(explains the overall status of patients at the start of the study),cell types of the tumor(cell type(1)-squamous cell cancer, cell type(2)-small cell cancer, cell type(3)-adenocarcinoma, cell type (4)-large cell carcinoma),treatment groups(standard test and chemotherapy test drug),status of the patients(0 represents censoring while 1 represents death),survival time(the time the observation period of the study commenced) and lastly their identification number of the patients.

## RESEARCH PURPOSE

I. To describe the notion about survival analysis and the variability between the diverse methods and test statistics used in analyzing survival data.
II. To determine whether there is a statistically remarkable difference between the survival distributions of the cell types, and also between the Standard test and Chemotherapy test treatment groups using the Veteran’s Administrative Lung cancer dataset.
III. To find out if they have any effect on the death of the male patients with advanced inoperable lung cancer.

Survival analysis was brought to existence to address the problem of right censoring. Right Censoring is the repeatedly experienced form of censored data and it can also be called **ADMINISTRATIVE CENSORING** where the observation interval ends without the event occurring as a result of lost to follow up, drop out or withdrawal (study termination).

There are three important methods used for survival analysis which include Actuarial (or Life table), Kaplan-Meier method and Cox-Regression.

### A) ACTUARIAL OR LIFE TABLE METHOD

This method takes into consideration diverse subjects who are censored and lost to follow up because they are alive at the termination of the analysis. These subjects happen to be alive at one time but showing different observation periods. It gives a simple, easy to know and easy to carry out method of analyzing survival data gotten at a specified time frame. Data are arranged in a tabular form within the fixed time set by the researcher for the study to be completed.

### B) KAPLAN-MEIER ORPRODUCT-LIMIT METHOD

This method makes use of the correct survival times (the failure time or censored time) instead of specified time frame to analyze survival. It processes the censored data very well and provides the test results of the survival probabilities and curves. Survival curves are not statistically distributed and are skewed to the right. Most clinical and epidemiologic studies prefer to use this method because it’s non-parametric nature and is arranged in a tabular form like Actuarial method. The survival function in Kaplan –Meier method is calculated as the number of deaths divided by subjects at risk.

### C) COX REGRESSION METHOD

This is a multivariate survival analysis that produces hazard ratios with both lower and upper 95% confidence intervals. Cox Regression approach is a semi-parametric procedure that can use both continuous and binary predictors. This method is used for designing the time to a particular event, established on the significance of the specified covariates. It gives the proportional hazard model, which can be drawn-out through the conditions of time-dependent covariates or conditions for strata. The proportional hazard model think that the covariates and the time to event are associated using this equation;

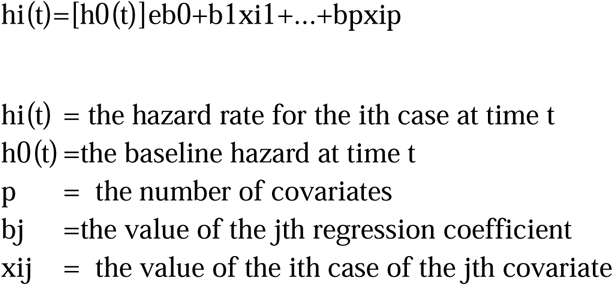

## SURVIVAL ANALYSIS TEST STATISTICS METHODS

### I) MANTEL-HAENSZEL OR LOG-RANK METHOD

The top most popular known firstly proposed by Mantel in 1966 followed by Cox in 1972. When the data includes censored data, this test compares both the actual and predicted number of failures and also set up a chi-square statistic to test the null hypothesis. It can only be used when the hazard rate between two groups are parallel (i.e. hazard ratio is constant).It is more acceptable for late entry data.

### II) GEHAN-BRESLOW-WILCOXON METHOD

This method is different from log rank method because the exponentiated coefficient [Exp(B)]of two groups must not be constant. It is more satisfactory for early failures.

### III) TARONE AND WARE METHOD

This test is more preferred than the above two methods because it focuses more on the failures that take place within the observation period instead of meeting the assumption of the proportional hazards. Also, it is can be applied for more than two groups and it works very well.

### IV) PRENTICE METHOD

This test is used when the assumption of the proportional hazard is violated and also focuses on early event times.

## EXPERIMENTS AND RESULTS

### LIFE TABLE RESULTS-METHOD 1

Life Table method was applied to the Veteran Administrative Lung Cancer dataset. The cell type variable in the real dataset was auto recoded into cell type 2(cell type 2(1)-adeno cell, cell type2 (2)-,large cell, cell type2(3)-small cell, cell type2(4)-squamous cell) in order to convert the categorical variable from string to numeric without losing any data. The survival time in days since the treatment was used as time variable, the status of the patient was used as the status variable where the single value used was 1 indicating event(number of death)and lastly, the celltype2 variable (four types of Lung cancer cell)served as the factor variable. Display time interval in days of 0 through the maximum time interval of type 999, and the displayed number of interval by type 1 was used.

**Figure 1.**
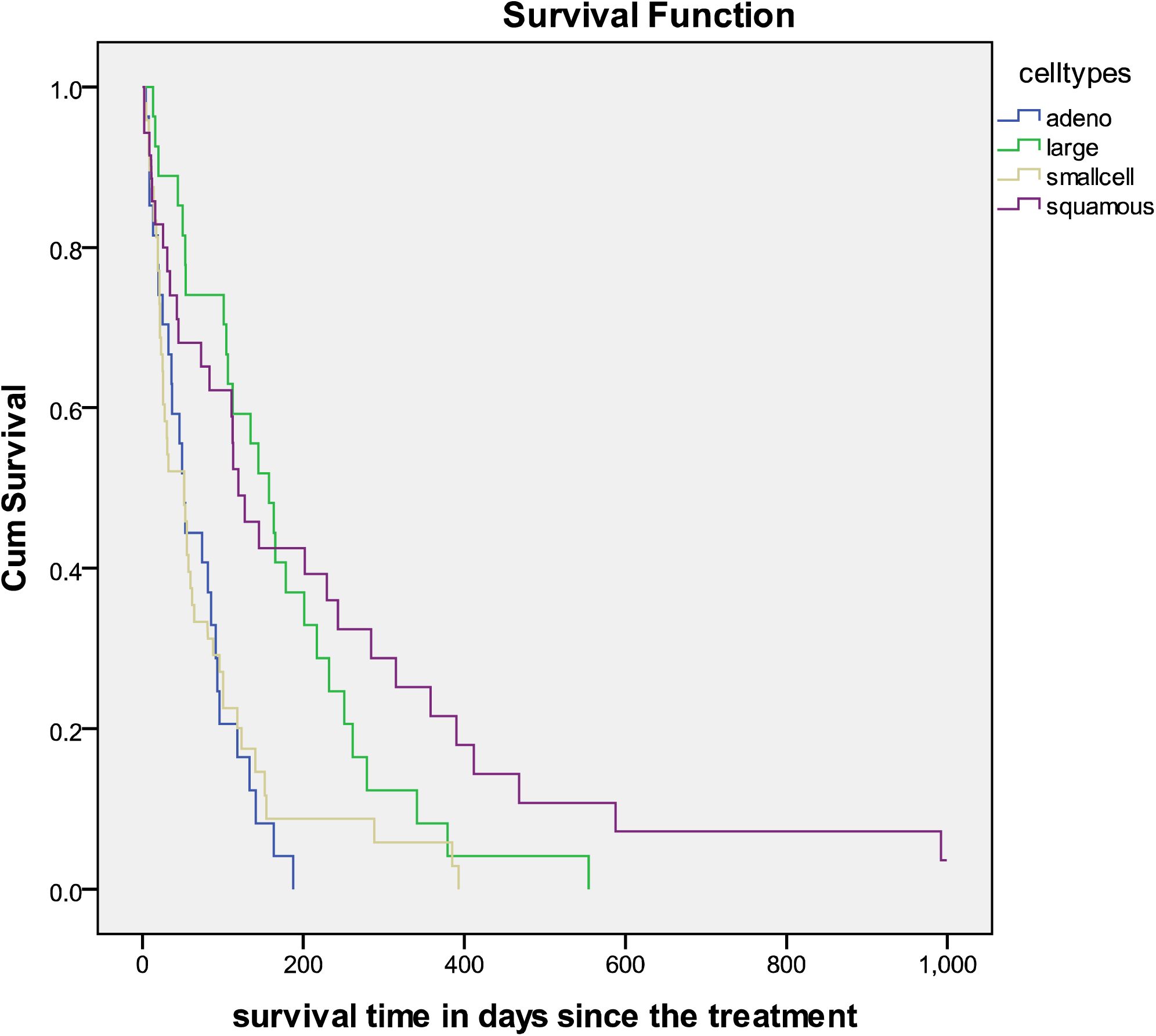
Survival Distribution of the different cell types.

The survival curves shows a picture of the life tables, the horizontal axis(x) represents the survival time in days after their treatment(time to event)t while the vertical axis(y) represents the survival probabilities. The adeno carcinoma cell type has the lowest survival curve succeeded by small cell lung cancer type, next is the large cell carcinoma type and lastly the squamous cell carcinoma type. We will be using the pairwise comparison table to explain if our observed differences happened by chance or not.

**TABLE 1:**
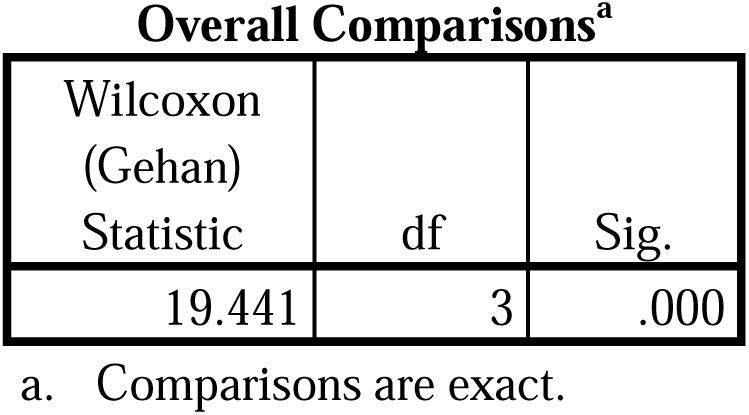
Comparisons for Control Variable: celltype2.

**TABLE 3:** PAIRWISE COMPARISON OF CELL TYPES2

**Pairwise Comparisons<sup>a</sup>**
| (I)<br>celltype<br>2 | (J)<br>celltype2 | Wilcoxon<br>(Gehan)<br>Statistic | df | Sig. |
| --- | --- | --- | --- | --- |
| 1 | dim 2 | 12.695 | 1 | .000 |
|  | ensi 3 | .052 | 1 | .820 |
|  | on1 4 | 5.827 | 1 | .016 |
| 2 | dim 1 | 12.695 | 1 | .000 |
|  | ensi 3 | 12.940 | 1 | .000 |
|  | on1 4 | .053 | 1 | .818 |
| 3 | dim 1 | .052 | 1 | .820 |
|  | ensi 2 | 12.940 | 1 | .000 |
|  | on1 4 | 7.742 | 1 | .005 |
| 4 | dim 1 | 5.827 | 1 | .016 |
|  | ensi 2 | .053 | 1 | .818 |
|  | on1 3 | 7.742 | 1 | .005 |

a. Comparisons are exact.

The above Wilcoxon (Gehan) statistic shows a p-value of 0.000 less than the 95% confidence level indicates that there is a statistical significance between the Lung cancer cell types. In this case, we reject the null hypothesis that says that all cell types are equal. From the pairwise comparison table we can deduct the following:

i. There is no statistical difference between adeno (1) and small cell type(3)–(lowest survival curves), and also between large cell(2) and squamous cell(4)-(lower survival curves).The p-value of the pairwise comparison in both group is greater than alpha significance level of 0.05 indicate that the survival curves are the same.
ii. Adeno celltype (1) is statistically different from large cell(2) and squamous cell(4) ;p-value <0.05 because it’s survival curve is lower than the two cell types.
iii. Small cell type(3) survival curve is lower than large cell(2) and squamous cell(4) ;p-value <0.05 because they are statistically different from the two cell types.

### KAPLAN MEIER RESULT -METHOD 2

Kaplan Meier method will be applied on the same dataset to determine if the Lung Cancer treatment survival distributions of one is better than the other, standard test or chemotherapy treatment (test drug).Survival time in days since after treatment was selected as the time variable, status of the patients with the event as 1 was used for the status variable while treatment group(categorical variable) was selected as the factor variable in order to examine the difference between the two groups. There are 69 inoperable males undergoing the standard test, 5 censored while 68 inoperable males used the test drug with 4 censored giving us a total of 137 participants and only 9 censored patients. The survival table, mean and median survival table and the survival plot was all plotted for a better understanding of our data.

**TABLE 4:**
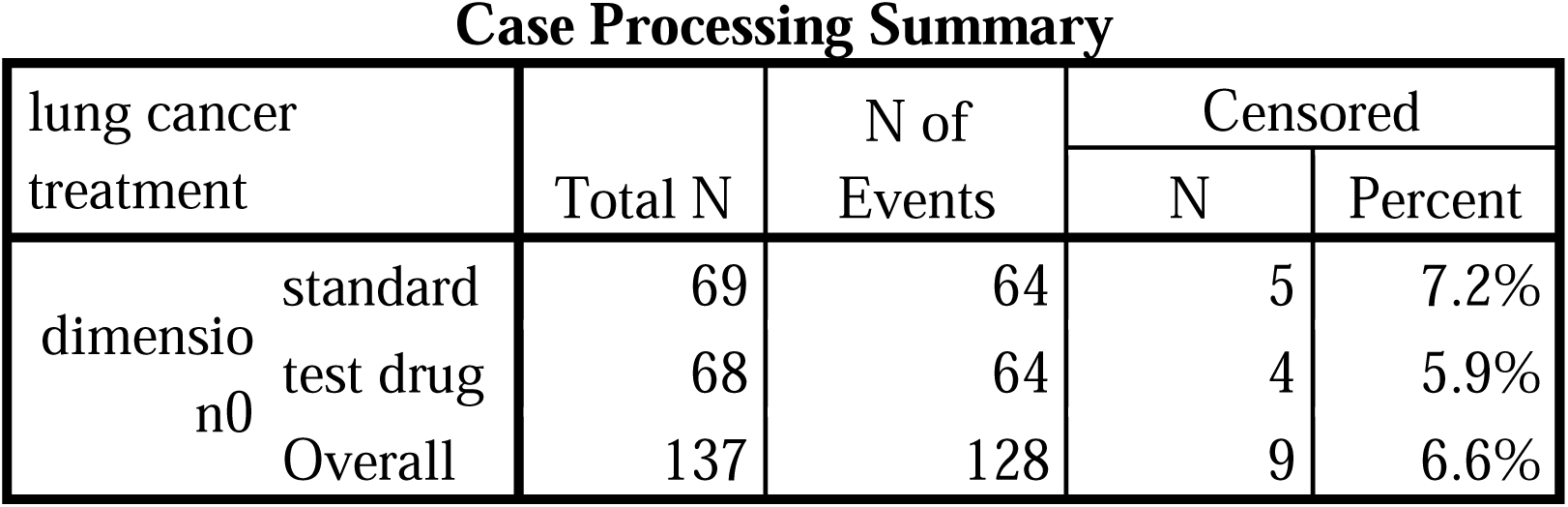
CASE PROCESSING SUMMARY OF LUNG CANCER TREATMENT

| Case Processing Summary |  |  |  |  |  |
| --- | --- | --- | --- | --- | --- |
| lung cancer treatment |  | Total N | N of Events | Censored |  |
|  |  |  |  | N | Percent |
| dimension0 | standard | 69 | 64 | 5 | 7.2% |
|  | test drug | 68 | 64 | 4 | 5.9% |
|  | Overall | 137 | 128 | 9 | 6.6% |

**TABLE 5:** MEANS AND MEDIANS FOR SURVIVAL TIME OF LUNG CANCER TREATMENT

| Means and Medians for Survival Time |  |  |  |  |  |  |  |  |
| --- | --- | --- | --- | --- | --- | --- | --- | --- |
| lung cancer treatment | Mean<sup>a</sup> |  |  |  | Median |  |  |  |
|  | Estimate | Std. Error | 95% Confidence Interval |  | Estimate | Std. Error | 95% Confidence Interval |  |
|  |  |  | Lower Bound | Upper Bound |  |  | Lower Bound | Upper Bound |
| standard | 123.928 | 14.961 | 94.605 | 153.251 | 103.000 | 19.810 | 64.173 | 141.827 |
| test drug | 142.061 | 27.023 | 89.097 | 195.026 | 52.000 | 14.202 | 24.164 | 79.836 |
| Overall | 132.777 | 15.368 | 102.655 | 162.898 | 80.000 | 15.721 | 49.187 | 110.813 |

a. Estimation is limited to the largest survival time if it is censored.

From the above table, the mean at 95% survival time for standard test is 123.928±14.961 while the mean survival time for test drug is 142.061±27.023 both are confidence interval. This result indicates that patients undergoing Chemotherapy treatment (test drug) has longer survival time than the ones undergoing standard test. Nevertheless, the authencity of this result can only be confirmed using the overall comparison table comprising the three test statistics.

**TABLE 6:** OVERALL COMPARISONS OF THE TEST STATISTICS

| Overall Comparisons |  |  |  |
| --- | --- | --- | --- |
|  | Chi-Square | df | Sig. |
| Log Rank (Mantel-Cox) | .008 | 1 | .928 |
| Breslow (Generalized Wilcoxon) | .961 | 1 | .327 |
| Tarone-Ware | .546 | 1 | .460 |

Test of equality of survival distributions for the different levels of lung cancer treatment.

Three statistics which includes Log rank, Breslow, and Tarone-Ware were selected for testing the equality of the survival distributions for the different levels of Lung Cancer Treatment and the pooled over strata was selected to specify the comparison. The above overall comparison table shows that Log rank has a p-value of 0.928, Breslow gave a p-value of 0.327 while Tarone-Ware gave a p-value of 0.460.This tells that all three p-values are greater than the alpha significance level of 0.05 which indicates that there is no statistically significant difference between the two Lung cancer treatment groups..

**FIGURE 2:**
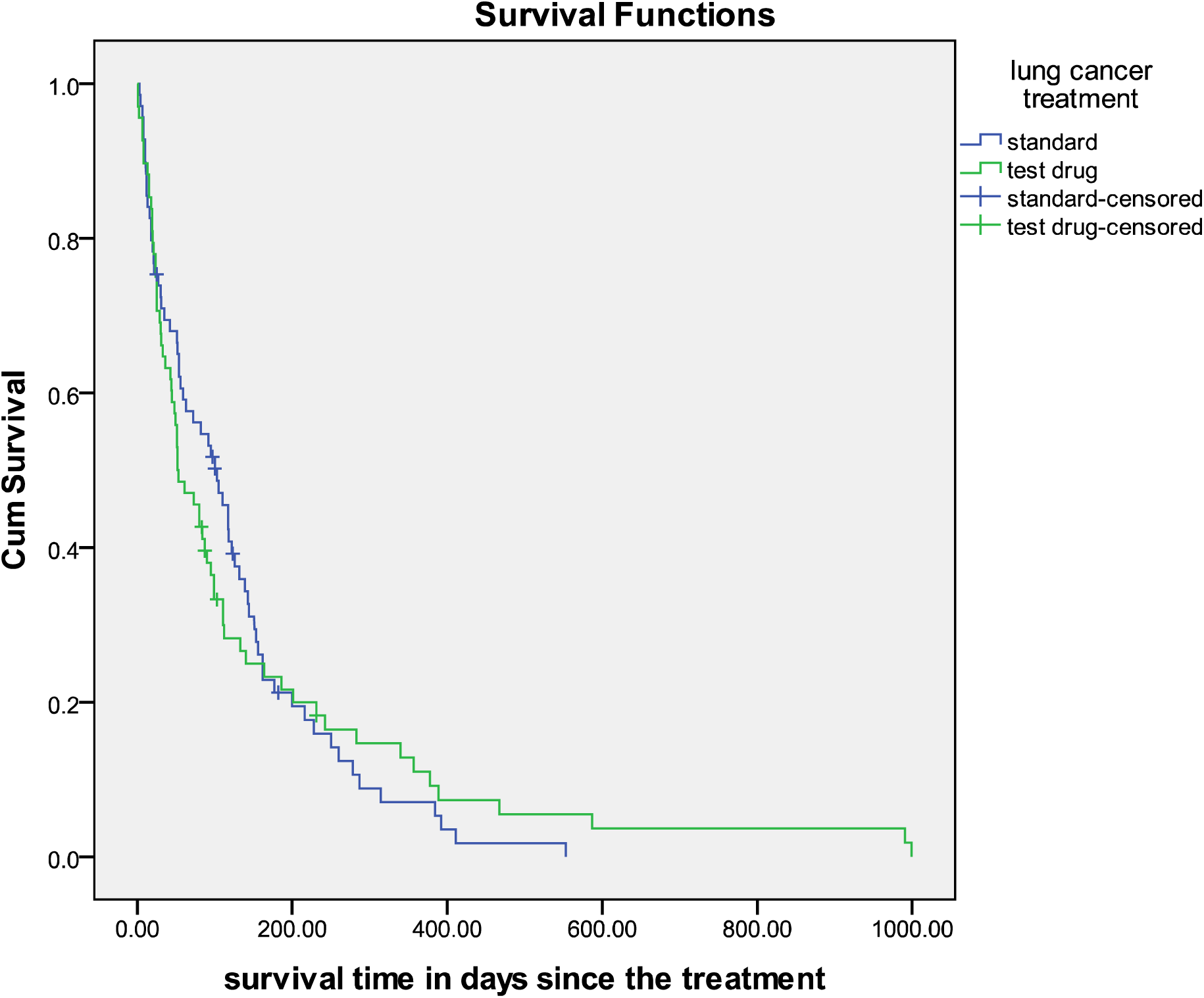
SURVIVAL DISTRIBUTION OF LUNG CANCER TREATMENT

The survival curve shows that at the beginning of the curve both treatments (standard and test drug) had a close death incident shape then in the middle of the curve the test drug decreased higher than the standard test and at the end of the curve it appeared that its survival time decreased lower when compared to the test drug that continued decreasing till the end. Howbeit, their different survival times are not statistically significant in relation to the overall comparison results of Log-rank, Breslow and Tarone-Ware test statistics.

### COX REGRESSION RESULT: METHOD 3

**TABLE 7:** OMNIBUS TESTS OF MODEL COEFFICIENTS

Omnibus Tests of Model Coefficients<sup>a</sup>
| -2 Log<br>Likelihood | Overall (score) |  |  | Change From Previous Step |  |  | Change From Previous Block |  |  |
| --- | --- | --- | --- | --- | --- | --- | --- | --- | --- |
|  | Chi-square | df | Sig. | Chi-square | df | Sig. | Chi-square | df | Sig. |
| 950.359 | 65.917 | 8 | .000 | 61.409 | 8 | .000 | 61.409 | 8 | .000 |

a. Beginning Block Number 1. Method = Enter

The above Omnibus tests of model coefficients gave a p-value of 0.000 which is less than the an alpha significance level of 0.05.This shows that the cell type2,prior therapy, age, diagnosis time, karnofsky score and treatment are statistically significant to the hazard events of the Lung cancer patients.

**TABLE 8:** REGRESSION COEFFICIENT AND P-VALUE TABLE

Variables in the Equation
|  | B | SE | Wald | df | Sig. | Exp(B) | 95.0% CI for<br>Exp(B) |  |
| --- | --- | --- | --- | --- | --- | --- | --- | --- |
|  |  |  |  |  |  |  | Lower | Upper |
| celltype2 |  |  | 17.916 | 3 | .000 |  |  |  |
| celltype2(1) | 1.188 | .301 | 15.610 | 1 | .000 | 3.281 | 1.820 | 5.917 |
| celltype2(2) | .400 | .283 | 1.999 | 1 | .157 | 1.491 | .857 | 2.595 |
| celltype2(3) | .856 | .275 | 9.687 | 1 | .002 | 2.355 | 1.373 | 4.038 |
| priortherapy | .007 | .023 | .097 | 1 | .755 | 1.007 | .962 | 1.054 |
| age | -.009 | .009 | .844 | 1 | .358 | .991 | .974 | 1.010 |
| diagtime | .000 | .009 | .000 | 1 | .992 | 1.000 | .982 | 1.018 |
| karno | -.033 | .006 | 35.112 | 1 | .000 | .968 | .958 | .978 |
| treatment | .290 | .207 | 1.958 | 1 | .162 | 1.336 | .890 | 2.006 |

### REGRESSION COEFFICIENT INTERPRETATION [EXP (β)]

The Exp (β) or Hazard Ratio for the first three celltypes2 of Lung cancer are relative to the fourth cell type2 (Squamous cell) known as the reference category. The hazard ratio for the cell type2(1),cell type2(2) and cell type2(3) are 3,281,1.491 and 2.355 times that of celltype2.Looking at the lower and upper 95% confidence level of the celltype2 they are not significant because their values are greater than 1,0.The regression coefficient of 1.005 for prior therapy shows that the male patients that had prior therapy have lesser risk of death than the males without any prior therapy, and their 95% confidence level indicates they are not significant. The age covariate Exp (B)has a coefficient of 0.991.The hazard ratio of the event(death occurrence) is reduced by 100%**-**(100% x 0.991) = 0.9% with each increase in Age, and their 95% confidence level indicates not significant. A unit increment in diagnosis time, increases the risk of death by 1.000, and the 95% confidence interval is not significant. The karnofsky score covariate regression coefficient is 0.968.The hazard ratio of the event is reduced by 100% - (100% x 0.968) = 3.2% with each increase in karnofsky score. The 95% confidence level (both lower and lower are <1.00) of karnofsky score is significant. The Exp(B) of 1.336 for treatment shows that the test drug have a lower risk of death than the standard test, and their 95% confidence level is not significant.

### THE ALPHA SIGNIFICANCE LEVEL INTERPRETATION

The celltype2, cell type (1), celltype(3) and karnofsky score covariates have p-value of 0.000, 0.000, 0.002 and 0.000 respectively. These p-values are lesser than the alpha significance level of 0.05 which shows that they are all statistically significant to the risk of death happening. While the other remaining covariates which include celltype2(2), prior therapy, age, diagnosis time and treatment are not statistically significant to death risk with p-values of 0.157, 0.755, 0.358, 0.992, and 0.162 respectively greater than 0.05 alpha significant level.

**FIG 3:**
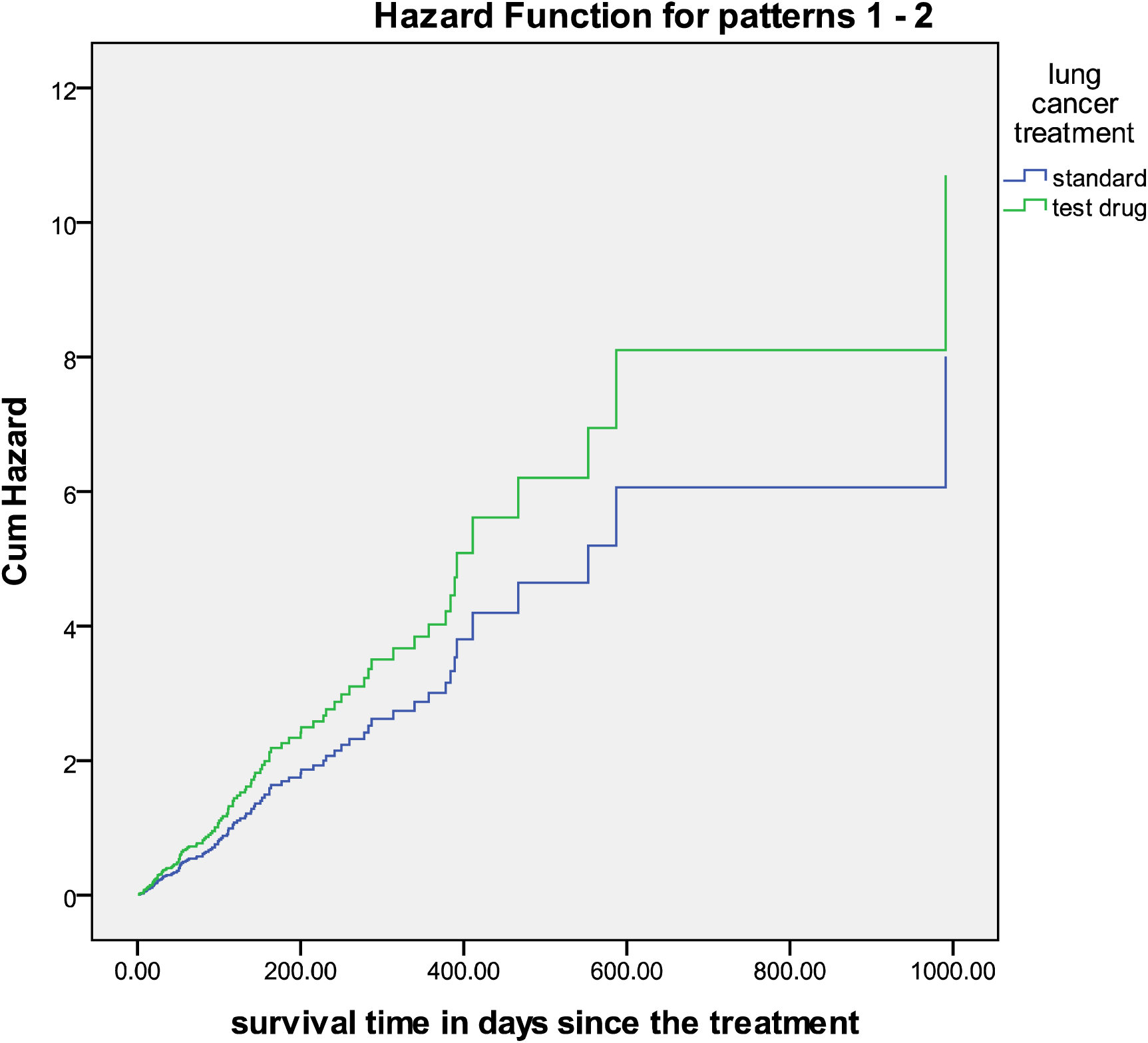
Hazard Function Graph of the two treatment group differences

## CONCLUSION

The Wilcoxon (Gehan) statistic from Life table method shows a p-value of 0.000 less than the 95% confidence level indicating that there is a statistical significance between the Lung cancer cell types. In this case, we reject the null hypothesis that says that all cell types are equal. The above overall comparison table from Kaplan Meier table shows that Log rank has a p-value of 0.928, Breslow gave a p-value of 0.327 while Tarone-Ware gave a p-value of 0.460.This tells that all three p-values are greater than the alpha significance level of 0.05 which indicates that there is no statistically significant difference between the two Lung cancer treatment groups. The mean at 95% survival time for standard test is **123.928±14.961** while the mean survival time for test drug is **142.061±27.023**.

Lastly, the Cox-Regression model showed that the squamous cell type2 (4), adeno cell type (1), small cell type (3) and karnofsky score covariates have p-values< 0.05 confidence level which indicates that they are all statistically significant to the risk of death happening. While the other remaining covariates which include large cell type2 (2), prior therapy, age, diagnosis time and treatment are not statistically significant to death risk of the lung cancer patients with p-value□0.05 confidence level. The treatment difference does not show any remarkable effect on the survival time of male patients with Lung Cancer.

## Data Availability

All data used for the study are available at http://lib.stat.cmu.edu/datasets/veteran

